# Cumulative Hardship: The Role of Perceived Injustice in Linking War-Related Trauma and Discrimination with Mental Health

**DOI:** 10.1101/2025.05.12.25327425

**Authors:** Christiane Wesarg-Menzel, Eileen Lashani, Mathilde Gallistl, Veronika Engert

## Abstract

Exposure to war-related trauma and post-migration discrimination are significant risk factors for mental health problems among refugees and migrants. Understanding the mechanisms linking these experiences to psychological outcomes is essential for informing targeted interventions in host countries. In the context of resettlement, this study examined whether perceptions of injustice and physiological stress regulation, measured through resting heart rate variability (HRV), mediated the relationship between trauma, perceived discrimination, and mental health problems. Data were collected from 98 refugees and migrants (21.4% female; *M*_age_ = 28.0 years) from Arabic-speaking countries living in Germany. Participants reported on pre-migration trauma, post-migration discrimination, perceived injustice, and depressive and post-traumatic stress disorder (PTSD) symptoms. A subsample of 67 participants underwent a 10-min electrocardiogram to assess resting HRV. Results revealed that pre-migration trauma was linked to increased PTSD symptoms, while post-migration discrimination was associated with higher depressive symptoms. Perceived injustice mediated the relationship between discrimination and mental health outcomes, with greater perceived discrimination being associated with higher perceived injustice, which in turn was related to both increased depressive and PTSD symptoms. These findings held after controlling for age and sex. No mediating effect of HRV was found, potentially due to limited statistical power. In conclusion, our study emphasizes the psychological burden of both pre- and post-migration stressors for refugees and migrants from Arabic-speaking countries. The mediation results suggest that addressing perceived injustice may be important for post-migration mental health interventions.

## Introduction

Individuals who have undergone migration or forced displacement are at increased risk for mental health problems (Kirmayer et al., 2011). Among newly arrived refugees and asylum seekers in Germany, pooled prevalence estimates range between 29.8–50.1% for depressive symptoms, and between 20.8–38.7% for Post-Traumatic Stress Disorder (PTSD) symptoms (Hoell et al., 2021). These rates by far exceed prevalence rates in the general German population (Bretschneider et al., 2017). Notably, particularly the comorbidity of depression and PTSD has been associated with longer-term psychosocial dysfunction among refugees (Momartin et al., 2004). In the current study, we investigated how two key risk factors—trauma exposure during pre-migration and ethnic discrimination during post-migration—contribute to mental health deterioration in refugees and migrants. Specifically, we examined their impact on depression and PTSD symptoms, and tested whether these associations would be mediated by perceptions of injustice and resting heart rate variability.

While trauma exposure is a known risk factor for mental health problems in refugee and migrant populations (Schlaudt et al., 2020; Wesarg-Menzel et al., 2024a), stressors often continue after settlement in a new country. Commonly reported post-migration stressors include ethnic discrimination, inadequate housing, unemployment, and worry about family remaining in the country of origin, all of which can further contribute to the decline in mental health (James et al., 2019; Li et al., 2016). Among these, perceived discrimination has garnered particular attention due to its strong association with depression among ethnic minority, migrant, and refugee populations (Giesebrecht et al., 2024; Ikram et al., 2016; Mewes et al., 215; Missinne & Bracke, 2012). Further evidence from cross-sectional and longitudinal studies implies perceived discrimination as a risk factor for PTSD (Brooks Holiday et al., 2020; Sibrava et al., 2019; Viazminsky et al., 2022).

Perceptions of injustice often emerge following exposure to traumatic events (Maercker & Horn, 2012). Research has shown that perceptions of injustice are likely to arise when individuals endure unnecessary suffering caused by others, experience irreparable loss, or face violation of their basic human rights (Sullivan, 2008), all of which are common in conflict settings. Persistent preoccupation with injustices of the past and present can play a relevant role in the maintenance or exacerbation of psychological symptoms (Silove, 1999; 2013). In accordance, strong links between perceived injustice and mental health problems were shown in post-conflict populations such as those residing in Timor-Leste (Rees et al., 2013; Silove et al., 2014; Tay et al., 2016) and West Papua (Rees & Silove, 2011). Yet, the literature focusing on the link between perceived injustice and mental health in populations exposed to conflict is still relatively scarce. In contrast, a substantially larger body of evidence exists for acute injury and chronic pain populations, where moderate to strong positive associations of perceived injustice with depressive and PTSD symptoms have been observed (for review, see Lynch et al., 2021; Pavilanis et al., 2023). As suggested by a prospective study, perceived injustice may represent a mechanism linking trauma to health outcomes: In patients admitted to a center for traumatic injuries (e.g., gunshot wounds and aggravated assaults), perceived injustice at 3 months post-injury mediated the relationship between trauma and elevated levels of depressive and PTSD symptoms at 6 months post-injury (Boals et al., 2020). As of yet, it remains unclear whether perceived injustice acts as a mechanism in the relationship between pre-migration exposure to trauma and mental health problems.

Perceived injustice may further link experiences of ethnic discrimination to mental health problems in individuals affected by migration. Support for this hypothesis stems from studies focusing on chronic back pain, showing that perceived injustice related to the pain condition mediated the association between perceived discrimination and depressive symptoms in a sample of Arab-Americans (Alnojeidi et al., 2025) and an ethnically diverse U.S. sample (Ziadni et a., 2020). Further, in a sample of African American and European American women, sexist events in the past year exerted a negative effect on depression, anxiety, and well-being, through a pathway of reduced just world beliefs and personal control. So far, the mediating role of perceived injustice in the relationship between ethnic discrimination and mental health problems has not been investigated in refugees and migrants.

Despite the repeatedly shown influences of trauma and discrimination on mental health (Giesebrecht et al., 2024; Schlaudt et al., 2020), the precise biological mechanisms underlying these relationships are not fully understood. One potential mechanism is the prolonged experience of stress and resulting dysregulation of biological systems involved in coordinating stress physiology (McEwen, 2008). Work on the physiological sequelae of extreme stress and trauma has placed particular focus on heart rate variability (HRV), which refers to the variation in the time intervals between consecutive heartbeats, known as inter-beat intervals (IBIs). As postulated in the Neurovisceral Integration Model (Thayer & Lane, 2000; 2009), HRV reflects the dynamic interplay between the autonomic nervous system and brain structures responsible for emotional and cognitive regulation. Specifically, HRV represents the capacity of the prefrontal cortex to exert top-down control over autonomic responses via the vagus nerve. Higher HRV indicates greater physiological flexibility, enabling adaptive responses to stress and environmental demands, while lower HRV has repeatedly been linked to stress, poor mental health, and chronic disease (Appelhans & Luecken, 2006; Smith et al., 2017; Thayer et al., 2012). Specifically, compelling evidence has revealed lower HRV in individuals with depression (Brown et al., 2018; Koch et al., 2019; Koenig et al., 2016) and PTSD (Ge et al., 2020; Schneider & Schwerdtfeger, 2020). HRV may not only be a marker, but also a precursor of resilience under stressful times. This is suggested by a recent study showing that higher HRV nearly two years prior to the COVID-19 pandemic predicted more functional emotion regulation strategies, as well as increased sense of safety and reduced worry in healthy adults during the lockdown (Makovac et al., 2021).

As a profound chronic stressor, perceived ethnic discrimination has rather consistently been linked to lower HRV (Panza et al., 2019). Yet, former evidence has primarily focused on African Americans (Hill et al., 2017; Hoggard et al., 2015; Utsey et al., 2007). No study has considered the relationship of discrimination and HRV among refugees. It has been suggested that experiences of discrimination elicit emotional distress to which the body responds with acute physiological changes, such as increases in heart rate and blood pressure, and the secretion of stress hormones (Clark et al., 1999; Sawyer et al., 2012). These physiological reactions may be prolonged by rumination about the perceived injustice (Brosschot et al., 2006; Gianferante et al., 2014; Silove, 2013). If exposure to discrimination occurs repeatedly, ultimately becoming chronic, this may lead to long-term dysregulation of physiological stress systems, giving rise to chronic reductions in HRV and deterioration of other stress system markers (Clark et al., 1999; Pascoe & Smart Richman, 2009). In partial support of this model, Hagen et al. (2021) found autonomic regulation (operationalized by an index including HRV and baroreflex sensitivity) to mediate the association between perceived ethnic discrimination and depressed mood in a multi-ethnic cohort including 9,492 participants from the Netherlands. However, this very modest mediating effect was attenuated after adjustment for socio-economic status. In another study conducted in Timor-Leste—a post-conflict country with a history of mass violence—age-related reductions in HRV emerged as a psychophysiological mechanism underlying enhanced vulnerability to distress and aggression following cumulative exposure to traumatic events (Liddell et al., 2016).

In summary, while significant evidence links trauma exposure and perceived discrimination to mental health problems, including symptoms of depression and PTSD, the underlying mechanisms remain inadequately understood. Relevant cognitive and biological mechanisms may be increased perceived injustice and reduced resting HRV. However, their roles in linking trauma and perceived discrimination to symptoms of depression and PTSD, particularly in refugee populations, has not been systematically explored. In the current study, we hypothesized that experiences of trauma and perceived discrimination would be related to higher levels of depressive and PTSD symptoms, with these associations being mediated by higher perceived injustice and lower HRV. This study is part of a broader project (Wesarg-Menzel et al., 2024a,b), and the specific hypotheses tested here were generated post hoc after data collection. Understanding the mechanisms linking stressor exposure to mental health is critical for refining interventions that support individuals who have undergone migration or forced displacement.

## Methods

### Procedure

Data stemmed from a larger study with refugees and migrants residing in and around Leipzig, Germany. The study was approved by the Ethics Board of the medical faculty of Leipzig University (ethics number: 405/18-ek). Participants were recruited from 2019–2023 through social media advertisements and flyers. Trained students conducted a structured telephone screening to determine participants’ eligibility. In a first lab visit lasting 2.5 hours, eligible participants filled out a range of self-report questionnaires and completed an empathy task (not subject to the current manuscript; findings reported in Wesarg-Menzel et al., 2024a). In a second visit lasting 4.5 hours, a subsample underwent an electrocardiogram (ECG) assessment during an empathic stress test, of which the 10 minutes baseline period is of interest to the current analyses. The study was performed in agreement with the Declaration of Helsinki. All participants gave their written informed consent and could withdraw from the study at any time. Participants received financial compensation for study participation.

### Participants

Ninety-eight participants (56 refugees and 42 migrants; 21.4% female; *M*_age_ = 28.0 years, *SD* = 4.8 years) took part in the study. The majority of participants (80.8%) came from Syria. Details regarding participant demographics and inclusion criteria can be found in prior publications (Wesarg-Menzel et al., 2024a,b) and in the Supplement. In short, participants needed to be aged between 20 to 40 years, come from a majority-Arabic-speaking country of origin, live in Germany for at least six months, speak Arabic as native language and German at an intermediate (B1) level, and have no diagnosed reading disability.

### Trauma

We employed the Arabic version of the trauma events section of the Harvard Trauma Questionnaire (HTQ; Mollica et al., 1992; Shoeb et al., 2007) to measure trauma exposure. Participants indicated whether they had experienced any of 42 traumatic events (e.g., oppression, imprisonment, combat exposure) before coming to Germany. A trauma score was computed by adding up the number of “yes” responses. Reliability was excellent (Cronbach’s alpha = .92).

### Perceived Ethnic Discrimination

Perceived ethnic discrimination was measured with the 22-item Perceived Ethnic Discrimination Questionnaire (PEDQ; Contrada et al., 2001). To this end, the PEDQ was translated into Arabic by a native, bilingual speaker and then back translated by a second bilingual individual to ensure linguistic equivalence (Brislin, 1986). Participants were asked to reflect on the past three months and report the frequency of specific events of discrimination, such as experiencing verbal rejection, unfair treatment, or aggression due to their ethnicity. Exemplary items are “How often has it been implied or suggested that because of your ethnicity you must be dishonest?” and “How often have you been exposed to offensive comments because of your ethnic group?”. Participants’ responses were captured on a Likert scale ranging from 1 (“never”) to 7 (“very often”), and aggregated to a mean score. Higher mean scores indicated higher perceived ethnic discrimination. Reliability was excellent (Cronbach’s alpha = .91).

### Perceived Injustice

Perceived injustice was assessed with the Injustice Experience Questionnaire (IEQ; Sullivan, 2008), translated into Arabic (forward and back translation method). The IEQ was originally designed to assess different thoughts concerning the sense of unfairness in relation to a musculoskeletal injury. It consists of 12 items related to two domains: irreparability of loss and blame/unfairness. Exemplary items include “It all seems so unfair.”, “Nothing will ever make up for all that I have gone through.”, and “Most people don’t understand how severe my condition is.”. For the current study, instructions were adapted to reflect how having to leave the home country and start a new life has affected participants. Items were scored from 0 (“never”) to 4 (“all the time”), and aggregated to a sum score, with higher scores indicating a greater degree of perceived injustice. In chronic pain samples, a cut-off score of 30 has been proposed to represent clinically relevant levels of perceived injustice (Sullivan, 2008). Reliability was good (Cronbach’s alpha = .86).

### Depressive Symptoms

The presence and severity of depressive symptoms were measured with the Beck Depression Inventory-II (BDI-II; Beck et al., 1996), translated into Arabic (forward and back translation method). The BDI-II contains 21 items that capture various cognitive, behavioral, and physiological symptoms associated with depression. Participants were asked to choose a phrase from a group of four response options (assigned scores from 0 to 3) that best reflected their experiences over the past two weeks. The total score ranges from 0 to 63, with higher scores indicating greater severity of depression. A score from 0 to 13 indicates minimal depression, 14 to 19 mild depression, 20 to 28 moderate depression, and 29 to 63 severe depression. Reliability was good (Cronbach’s alpha = .84).

### PTSD Symptoms

We measured PTSD symptom severity with the trauma symptom section of the Arabic version of the HTQ (Mollica et al., 1992; Shoeb et al., 2007). Participants indicated the degree to which they were distressed in the past week by trauma symptoms such as experiencing “recurrent nightmares”, being “unable to feel emotions”, or “feeling irritable or having outbursts of anger” on 45 items, using a Likert scale ranging from 1 (“not at all”) to 4 (“extremely”). The current study makes use of the first 16 items of the trauma symptom section, which correspond to symptoms of PTSD according to the Diagnostic and Statistical Manual of Mental Disorders (DSM)-IV (American Psychiatric Association, 1994). Higher mean scores reflect more severe PTSD symptoms. The HTQ manual recommends a cut-off score of 2.5 to identify clinically significant levels of PTSD (Mollica et al., 1992). Reliability was good (Cronbach’s alpha = .87).

### Resting Heart Rate Variability

Data on resting HRV was gathered in a subsample of 67 participants (56 males) with a Zephyr Bioharness 3 chest belt (Zephyr Technology, Annapolis, Maryland, USA), which recorded a continuous ECG at a frequency of 250 Hz. Participants were instructed to sit still and relax during a resting period of 10 min. After data collection, a trained student manually corrected artifacts in the ECG raw data using python-based in-house software. Two additional students re-checked all corrections made. The 10-min ECG recordings were split into two 5-min timeframes, for which averages for HRV (specifically the Root Mean Square of Successive Differences [RMSSD]) were calculated per participant using the python package “hrv-analysis” (Champseix et al., 2021). To ensure data quality, we excluded timeframes in which more than 10% of the recording had to be cut.

### Data Analysis

#### Data Processing and Missing Data

Statistical analyses were performed in R (version 4.4.0; R Core Team 2024). The data set and analysis code are publicly available at https://osf.io/ukh5f/?view_only=2b3e9386aee3440b884a97ede6dcb1a6.

HRV data were log-transformed to approach normal distribution. No outliers beyond three standard deviations below or above the group mean were detected. Due to technical issues, HRV data was missing from two participants. In addition, HRV data quality was too low (i.e., >10% needed to be cut) in one participant. Thus, data on resting HRV was missing for three participants overall.

Missing questionnaire data was handled depending on their respective scoring. For the HTQ trauma score, three participants were missing one item each. Missing items were treated as non-affirmative responses and therefore not added to the sum score. For the HTQ PTSD and PEDQ mean scores, five participants had up to two missing items. In these cases, the means of the available items were calculated for each participant. IEQ and BDI scores had no missing data.

#### Statistical Analysis

Our statistical analysis followed a data-driven approach. Using structural equation modeling (SEM), we successively mapped the data onto our conceptual model. As a starting point, a direct effects model was tested where trauma and discrimination predicted symptoms of PTSD and depression (Model 1). Covariances were allowed between the two predictors and the two outcomes. We then proceeded to add perceived injustice (Model 2a) and HRV (Model 2b) as central mediators. Since HRV data were only available for a subset of the sample (*n* = 64), statistical power was insufficient to test a combined model including both mediators. In each step, weak paths were removed until optimal model fit was reached. This way, the resulting models represent the best possible adjustment of the conceptual model to fit the observed data. Additionally, we conducted two covariate analyses for each model. First, we controlled for the effects of age and self-reported sex. Second, we controlled for potential differences between refugees and migrants in our sample.

To evaluate the models’ goodness of fit, we computed Pearson’s chi-squared test (χ^2^), the Comparative Fit Index (CFI), Tucker-Lewis Index (TLI), and the Root Mean Square Error of Approximation (RMSEA) with its 90 % confidence interval. A model was considered to have an acceptable fit if the chi-squared test was non-significant (*p* < .05), CFI and TLI exceeded 0.90, and RMSEA was below 0.08. For all other analyses, two-sided *p*-values < .05 were considered statistically significant.

All models were implemented using maximum likelihood estimation in the lavaan package for R (version 0.6-18, Rosseel, 2012). To ensure robustness of our results, estimates were computed with 1,000 bootstrapped iterations. Multicollinearity was probed for and found unproblematic with all VIFs of the largest model assuming values below five.

## Results

Descriptive statistics and correlations of all variables used in this study can be found in supplementary section S1. On average, participants reported having experienced 13 out of 42 traumatic events listed in the HTQ, reflecting high levels of trauma exposure. Across all items of the PEDQ, participants indicated relatively infrequent experiences of ethnic discrimination (i.e., a mean score of 2.4, which falls between the “never” and “sometimes” categories).

Eleven of the 98 participants (11.22%) reported feelings of injustice that surpassed the clinical relevance threshold proposed based on data from chronic pain samples. An overview of the number of participants with clinically significant levels of depression and PTSD is presented in supplementary section S2. Overall, our sample reported rather low levels of depression and PTSD with over 90% reporting only minimal to mild symptoms on the BDI as well as sub-clinical levels on the HTQ.

### Model 1: Direct Effects of Trauma and Discrimination on PTSD and Depression

In the first model, we explored the relationships between trauma, perceived discrimination, and symptoms of PTSD and depression. The results supported a significant association between trauma and PTSD symptoms, with higher levels of trauma predicting greater PTSD symptom load. On the other hand, perceived discrimination significantly predicted depressive symptoms, indicating that higher levels of perceived discrimination were associated with more severe depressive symptoms. No significant associations were found between trauma and depression or between discrimination and PTSD, leading to the removal of these paths from the model. Additionally, the predictors showed moderate covariance, and the outcomes exhibited a strong correlation. The final path model is presented in Figure 1, with full statistical details summarized in Table 1. Overall, the model demonstrated good fit: χ^2^(2) = 2.67, *p* = .263, CFI = 0.990, TLI = 0.969, RMSEA = 0.059 (90% CI [0.00, 0.218]).

**Figure 1.**
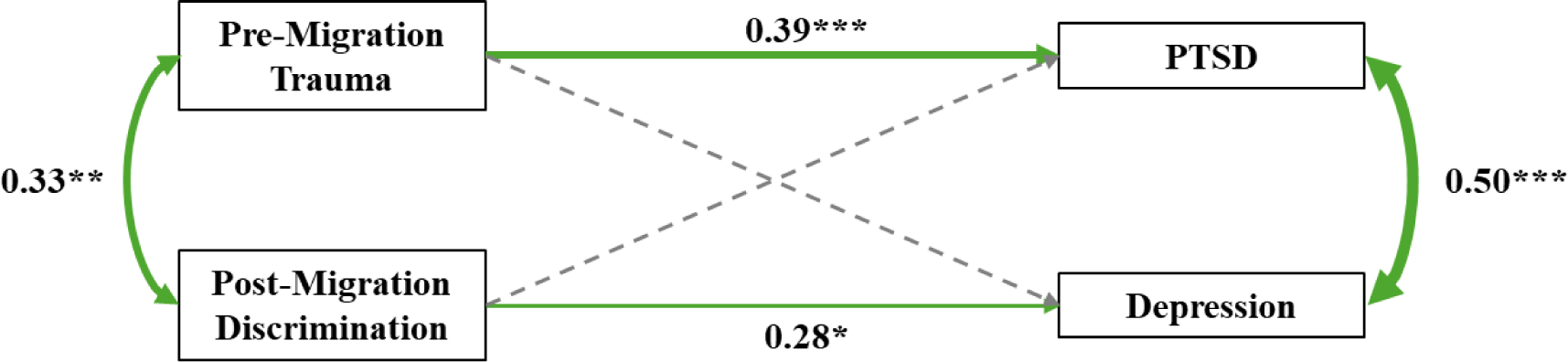
Model 1. *Note.* The term “migration” refers to both forced migration (refugees) and voluntary migration (migrants). PTSD refers to Post-Traumatic Stress Disorder. Green paths reflect positive associations. Grey dashed paths represent paths that were removed from the hypothesized model to enhance model fit. \**p* < .05; \*\**p* < .01; \*\*\**p* < .001.

**Table 1.**
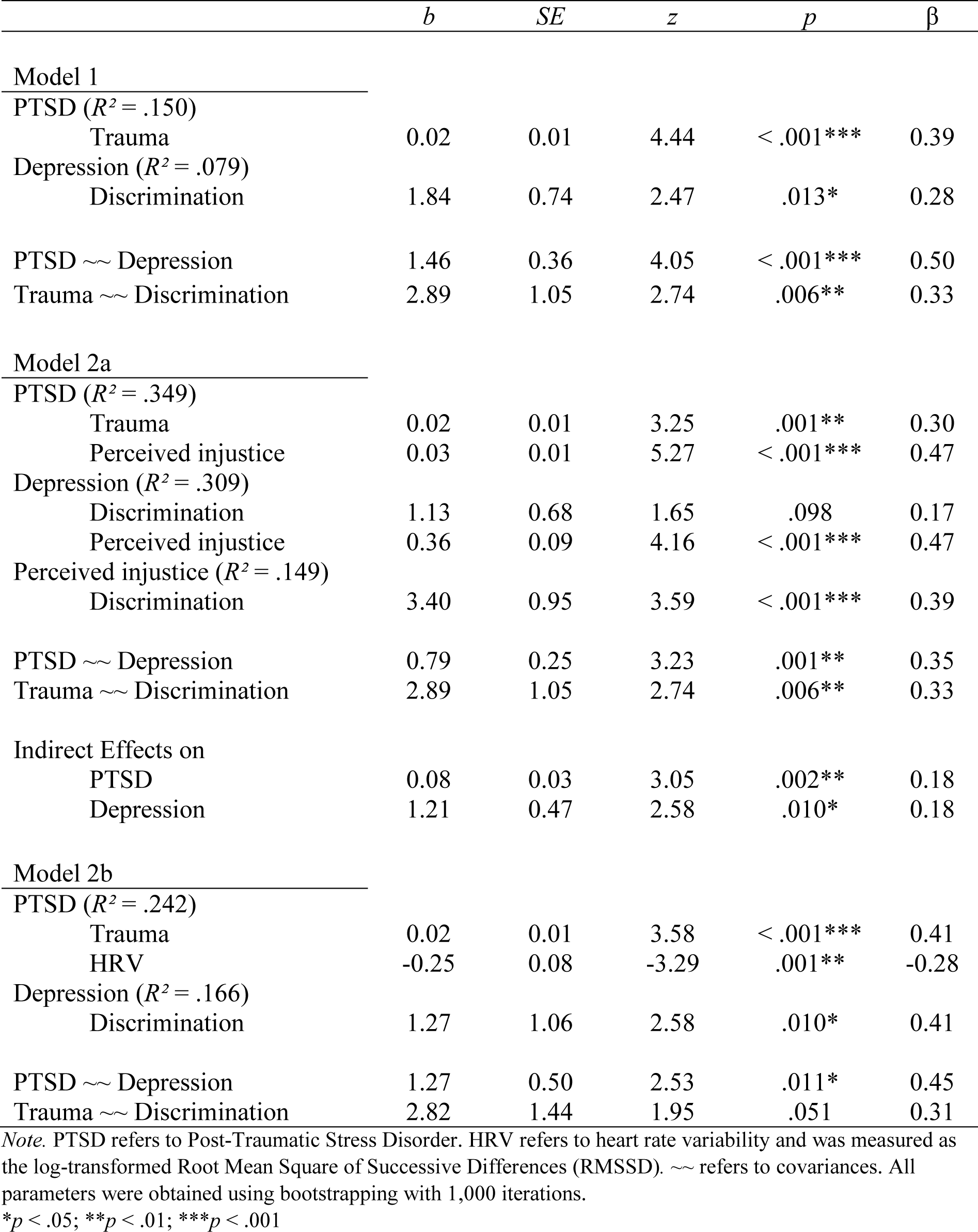
Regression, Covariance, and Mediation Coefficients of Models 1, 2a, and 2b.

### Model 2a: Mediation Through Perceived Injustice

In Model 2a, we extended our analysis to examine whether the relationship between trauma, discrimination, and mental health outcomes was mediated by perceived injustice. Trauma remained a significant predictor of PTSD symptoms but was not significantly linked to perceived injustice, leading to the removal of this path. Consequently, no mediation of trauma effects on PTSD via perceived injustice was identified. However, perceived discrimination significantly predicted feelings of injustice, which, in turn, had moderate to strong effects on both PTSD and depressive symptoms. Indirect effects were significant for both outcomes, suggesting that beyond the direct effect of trauma on PTSD, perceived discrimination contributed to poorer mental health through heightened feelings of injustice. Notably, the mediation effect on depression outweighed the direct effect of discrimination, rendering the latter non-significant in our sample. The predictors and outcomes showed moderate covariance. The final model, depicted in Figure 2, demonstrated excellent fit: χ^2^(3) = 3.51, *p* = .320, CFI = 0.996, TLI = 0.985, RMSEA = 0.042 (90% CI [0.00, 0.180]). Full parameter estimates are presented in Table 1.

**Figure 2.**
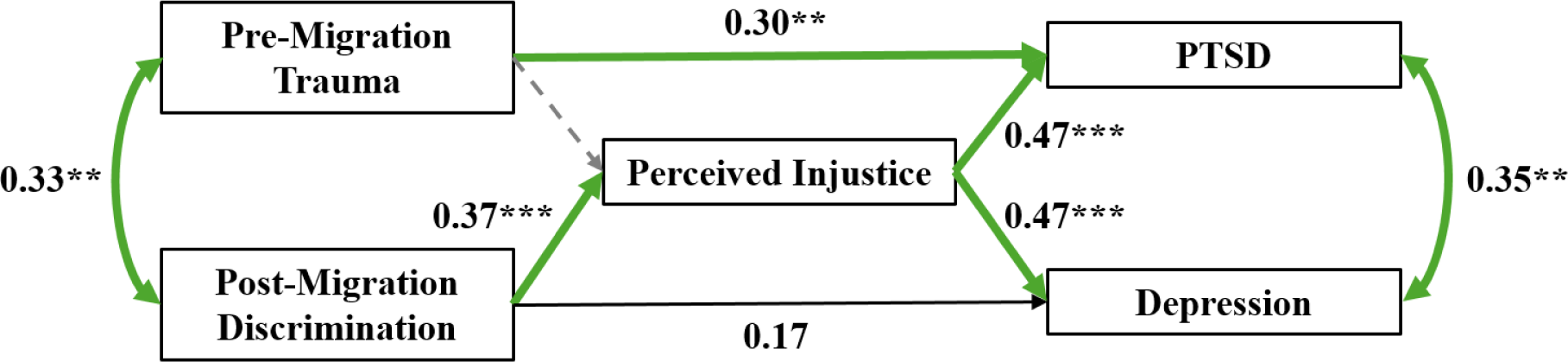
Model 2a. *Note.* The term “migration” refers to both forced migration (refugees) and voluntary migration (migrants). PTSD refers to Post-Traumatic Stress Disorder. Green paths reflect positive associations. Black paths reflect non-significant associations. Grey dashed paths represent paths that were removed from the hypothesized model to enhance model fit. \*\**p* < .01; \*\*\**p* < .001.

### Model 2b: Mediation Through HRV

In Model 2b, we introduced resting HRV (in terms of log-transformed RMSSD) as a mediator, following the same structure as in Model 2a. Due to missing ECG data, this model included only 64 out of the 98 participants, limiting statistical power. Trauma remained a significant predictor of PTSD symptoms, and discrimination significantly predicted depressive symptoms. However, neither trauma, perceived discrimination, nor depressive symptoms were significantly associated with HRV, leading to the removal of these paths and precluding further mediation analysis. What could be observed was a negative direct effect of HRV on PTSD symptom severity, suggesting a potential buffering role of parasympathetic regulation. Significant covariance was observed between the mental health outcomes but remained at a marginal level for the predictors. The model demonstrated good fit: χ^2^(5) = 6.14, *p* = .293, CFI = 0.978, TLI = 0.957, RMSEA = 0.060 (90% CI [0.00, 0.191]). The final path model is illustrated in Figure 3, and complete parameter estimates are presented in Table 1.

**Figure 3.**
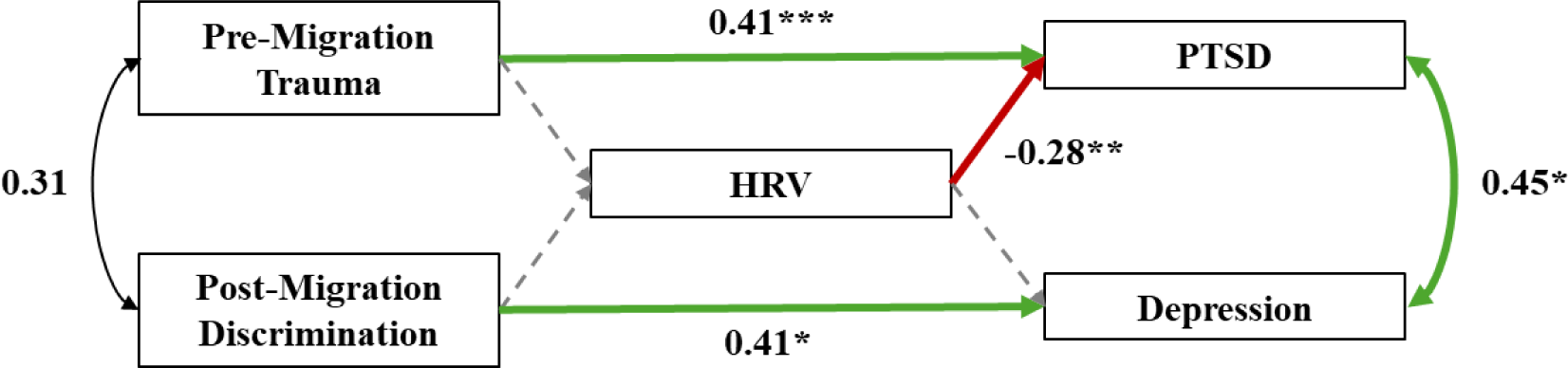
Model 2b. *Note.* The term “migration” refers to both forced migration (refugees) and voluntary migration (migrants). HRV refers to heart rate variability and was measured as the log-transformed Root Mean Square of Successive Differences (RMSSD). PTSD refers to Post-Traumatic Stress Disorder. Green paths reflect positive associations. Red paths reflect negative associations. Black paths reflect non-significant associations. Grey dashed paths represent paths that were removed from the hypothesized model to enhance model fit. \**p* < .05; \*\**p* < .01; \*\*\**p* < .001.

### Covariate Analyses of Age, Sex, and Refugee or Migrant Status

The first covariate analysis, which controlled for age and sex effects in our main models, is fully reported in Supplementary Section S3. Female sex was significantly associated with higher PTSD symptoms, older age with higher depressive symptoms, and both with greater perceived injustice. However, including these covariates resulted in poor model fit, likely due to power limitations. Importantly, their inclusion did not alter the pattern of results for our variables of interest.

The second covariate analysis, accounting for potential differences between refugees and migrants, is detailed in Supplementary Section S4. Belonging to either group did not significantly predict symptoms of PTSD or depression. However, a small effect was observed for perceived injustice, with refugees reporting higher levels of perceived injustice. The primary findings remained largely consistent, although the association between discrimination and depressive symptoms in Model 2a weakened and lost significance. Notably, the inclusion of the covariate again resulted in poor model fit, warranting cautious interpretation of these results.

## Discussion

In a sample of refugees and migrants from Arabic-speaking countries residing in Germany, we examined the associations between pre- and post-migration stressors and mental health problems, and tested whether perceived injustice and HRV mediated these relationships. Results indicated that pre-migration trauma was significantly associated with increased PTSD symptoms, while post-migration discrimination was significantly linked to elevated depressive symptoms. Perceived injustice mediated the association between discrimination and mental health outcomes, such that greater perceived discrimination was associated with more negative appraisals of injustice, which in turn were related to both increased depressive and PTSD symptoms. In a subsample of participants with available physiological data, no evidence was found for a mediating role of HRV in the associations between stressors and mental health problems, potentially due to limited statistical power. However, higher HRV was associated with lower PTSD symptoms. Results remained stable in covariate analyses controlling for the influences of age and sex, as well as for potential differences between refugee and migrant populations.

The specificity observed in the associations between war-related trauma and PTSD, as well as between ethnic discrimination and depression, aligns with established etiological models. According to the Diagnostic and Statistical Manual of Mental Disorders (DSM)-5 (American Psychiatric Association [APA], 2013), directly experiencing or witnessing a traumatic event, or learning that a traumatic event occurred to a close person, is a prerequisite for PTSD. Consistent with our findings, extensive research supports that exposure to war-related trauma is a primary risk factor for PTSD, both in civilians (Johnson & Thompson, 2008) and military personnel (Dohrenwend et al., 2013). Despite the high levels of war-related trauma reported in our sample, only a few participants exhibited clinically significant levels of PTSD. This may reflect resilience among our participants, which could at least in part stem from a selection bias, as those with better mental health may feel more capable of participating in a study.

In contrast to PTSD, depressive disorders are defined in the DSM-5 (APA, 2013) irrespective of their cause. Although our participants, on average, reported experiencing ethnic discrimination rather rarely, a link to depressive symptoms was nonetheless observed, with symptoms ranging predominantly in the mild range. This finding aligns with previous cross-sectional and longitudinal studies showing that perceived ethnic discrimination is a predictor of depressive symptoms (Schulz et al., 2006; Wang & Narcisse, 2025).

We hypothesize that with a larger sample and greater statistical power, a significant association between war-related trauma and depressive symptoms, as well as between ethnic discrimination and PTSD symptoms, would emerge. The following considerations lend support to this hypothesis: First, trauma has been conceptualized as a general vulnerability factor for psychopathology (Hogg et al., 2023). Second, while discrimination is generally considered an extreme chronic stressor rather than a traumatic event per se, some forms of discrimination, particularly those involving threats to life, are inherently traumatic. In line with these considerations, exposure to war-related trauma has been found to increase the risk of depressive disorders (Bonde et al., 2018; Ahmed et al., 2024), and ethnic discrimination has been prospectively linked to PTSD symptoms (Bird et al., 2021). Third, depressive and PTSD symptoms were moderately correlated in our sample, suggesting potential symptom overlap, mutual influence, or shared underlying causes.

As a cognitive mechanism, we examined whether perceived injustice mediated the relationship between pre- and post-migration stressors and mental health problems. Perceived injustice was positively associated with post-migration discrimination, but showed no significant relationship with pre-migration trauma. Notably, perceived injustice fully mediated the association between discrimination and depressive symptoms, rendering the direct effect of discrimination on depressive symptoms non-significant. Further, although there was no direct association between discrimination and PTSD symptoms, perceived injustice was positively related to both discrimination and PTSD symptoms, indicating an indirect-only mediation effect. Consistent with previous findings in post-conflict populations (Rees et al., 2013; Silove et al., 2014; Tay et al., 2016), these results highlight the important role of perceived injustice in shaping mental health outcomes among post-migration populations. While discrimination increases vulnerability to mental health problems, the individual’s processing and appraisal of the migration experience appear to be critical (Kuo, 2014; Lazarus and Folkman, 1984). In particular, persistent rumination about the perceived unfairness of one’s situation and the irreparability of losses may heighten emotional distress (Borders & Liang, 2011). This process may contribute to the development of various mental health problems, including depressive and PTSD symptoms, as observed in our sample. As suggested by our findings, interventions aimed at reducing perceived injustice may mitigate the negative mental health outcomes associated with discrimination.

Apart from perceived injustice, other mechanisms likely play a role in increasing mental health risk after discrimination. On a psychological level, experiences of discrimination may erode self-esteem, foster internalized negative beliefs, and contribute to a diminished sense of social belonging, all of which are known risk factors for depression (Espinosa 2020; Freire & Hurd, 2023; Quinn et al., 2015). Moreover, discrimination often leads to heightened vigilance for rejection, emotion dysregulation, and rumination, which can further exacerbate depressive symptoms (Bernard et al., 2022; Himmelstein et al., 2015; Mekawi et al., 2020). On a social level, individuals who experience discrimination may withdraw from interpersonal relationships or perceive lower social support, thereby reducing a critical buffer against stress (Shim et al., 2012). In the context of migration, these psychological and social mediators may already be compromised by pre-migration trauma exposure and the migration-related disruption of close relationships. When discrimination is added to these pre-existing challenges, it creates a double burden that can significantly aggravate health risk. In this regard, the positive covariance observed between trauma exposure and discrimination in our sample is worth highlighting. This finding suggests that refugees and migrants who have experienced more trauma may also be more likely to perceive ethnic discrimination, which may relate to greater vulnerability or threat sensitivity to racism (see Matheson et al., 2019).

Interestingly, feelings of injustice were associated with perceived discrimination rather than war-related trauma, which may be explained by several factors. First, while war-related traumatic experiences in the country of origin occurred longer time ago, ethnic discrimination may unfold as a recurrent experience in the daily lives of refugees and migrants. Additionally, discrimination may be perceived as a targeted and personal violation, with individuals feeling singled out due to their ethnic identity (Contrada et al., 2000). This directness can evoke a strong sense of unfairness and moral transgression, which aligns closely with the construct of perceived injustice (Sullivan, 2008). In contrast, war-related trauma is often attributed to broader, less controllable geopolitical forces (Gartzke, 1999; Muldoon, 2024). Consequently, while war-related trauma can have profound psychological effects, it may evoke perceptions of injustice to a lesser extent than post-migration discrimination.

As a biological mechanism, we tested whether resting HRV mediated the associations between pre- and post-migration stressors and mental health problems. Due to ECG data being available for only a subsample of 64 participants, findings should be interpreted with caution. Neither trauma nor discrimination was significantly associated with HRV, which precluded further mediation analyses. Previous research has suggested that the relationship between adversity exposure and HRV may be more pronounced in clinical populations (Sigrist et al., 2021; Wesarg et al., 2022). This may indicate that alterations in physiological stress systems, as indexed by HRV, emerge primarily in individuals more severely affected by pre- and post-migration stressors, and who consequently develop psychopathology (Wesarg et al., 2022). The absence of a significant association between stressor exposure and HRV in our study may therefore relate to the fact that our participants reported relatively low mental health problems, suggesting resilience at both the physiological and psychological level. Notably, however, we did observe a significant association between lower HRV and higher PTSD symptoms. This finding is consistent with prior evidence identifying reduced HRV as a potential endophenotype of PTSD (Ge et al., 2020; Schneider & Schwerdtfeger, 2020), possibly reflecting diminished flexibility to cope with environmental challenges (Thayer & Lane, 2000; 2009).

This study has several limitations that warrant consideration. First, the research question was formulated after data collection, and hence, analyses were not pre-registered. Second, the overall sample size was relatively small, particularly for analyses involving HRV data, which limited statistical power. Third, the sample included a disproportionately low number of female participants and was restricted to individuals in early and middle adulthood, limiting the generalizability of our findings across gender and age groups. Fourth, we did not assess the timing or duration of participants’ exposure to trauma and discrimination, which may have influenced observed associations among stressor exposure, perceived injustice, HRV, and mental health outcomes. Fifth, we relied exclusively on participants’ subjective perceptions of discrimination, without collecting objective data on actual exposure. As a result, the extent to which discrimination objectively occurred remains unknown. Importantly, psychological biases can lead individuals to either downplay clear instances of discrimination or, conversely, to exhibit hypervigilance to ambiguous cues that might constitute discrimination (Lewis et al., 2015). Nonetheless, focusing on subjective perceptions is justified, as the appraisal of stressors plays a central role in the development of stress-related symptoms (Anisman, 2014). Finally, the cross-sectional nature of the study precludes any inference of temporal or causal relationships. Thus, we cannot rule out, for instance, that mental health problems preceded exposure to war-related trauma or discrimination.

In conclusion, our study highlights the significant psychological burden associated with both pre- and post-migration stressors among refugees and migrants from Arabic-speaking countries residing in Germany. While war-related trauma was associated with PTSD symptoms, perceived ethnic discrimination was linked to depressive symptoms. Further, perceived injustice emerged as a significant mediator of the relationship between discrimination and mental health problems. These findings underscore the importance of addressing perceived injustice in post-migration mental health interventions.

## Acknowledgements

We cordially thank Beyond Conflict for their conceptual input and financial support of the study. We further thank Elisabeth Murzik and Sylvia Tydecks for their involvement in study organization and coordination, and Henrik Grunert for technical assistance. We also thank our participants and all students of the Social Stress and Family Health Research Group at the Max Planck Institute for Human Cognitive and Brain Sciences involved in recruitment and data collection.

## Data Availability Statement

The data set and analysis code used in the present study are publicly available at https://osf.io/ukh5f/?view_only=2b3e9386aee3440b884a97ede6dcb1a6

