## Supplementary material for "Cumulative Hardship: The Role of Perceived Injustice in Linking War-Related Trauma and Discrimination with Mental Health": Supplemenzt

*Submitted to Journal of Neural Transmission*

Christiane Wesarg-Menzel, Eileen Lashani, Mathilde Gallistl, & Veronika Engert

### ~ Supplementary Materials ~

#### Detailed Information on Participants

Based on definitions provided by UNHCR (2023), individuals who were forced to flee their home countries due to conflict or persecution were classified as refugees. Our operationalization of the refugee group also included asylum seekers—individuals who had not yet received legal recognition as refugees and were still awaiting a decision on their asylum application (UNHCR, 2023). In contrast, individuals who voluntarily left their home countries to work, study, or reunite with family members were classified as migrants. These two groups were selected to examine potential differences in empathic abilities between individuals with war-related trauma (refugees) and those without such trauma but with comparable cultural backgrounds (migrants).

Refugees were eligible for inclusion if they had experienced war-related trauma (i.e., they had fled war, violence, conflict, or persecution). Exclusion criteria for the refugee group were exposure to any major non-war-related trauma (e.g., maltreatment, severe accidents, natural disasters); diagnosis of a psychiatric disorder within the past two years, with the exception of PTSD and depression; and the presence of severe depressive symptoms within the past four weeks, defined as a score of $\geq 5$ on the depression section of the Structured Clinical Interview for DSM-IV Personality Disorders (SCID; First et al., 1996). Migrants were eligible for inclusion if they had never experienced major trauma (including war-related trauma) and had not been diagnosed with any psychiatric disorder within the past two years.

Group allocation was initially based on a telephone screening conducted with all participants. However, subsequent analysis of responses to the more detailed Harvard Trauma Questionnaire (HTQ; Mollica et al., 1992) revealed that a substantial proportion of migrants had also been exposed to significant war-related trauma.

### S1 Scale Correlations

| | *M* | *SD* | 1 | 2 | 3 | 4 | 5 | 6 | 7 |
|---|---|---|---|---|---|---|---|---|---|
| 1. Sex (female) | | | | | | | | | |
| 2. Age | 28.03 | 4.84 | -.19 | | | | | | |
| 3. Trauma (HTQ) | 12.70 | 8.45 | -.28** | .12 | | | | | |
| 4. Discrimination (PEDQ) | 2.43 | 1.05 | -.07 | -.11 | .33** | | | | |
| 5. Perceived injustice (IEQ) | 18.54 | 9.25 | .24* | .08 | .28** | .39** | | | |
| 6. Depression (BDI) | 8.90 | 7.03 | .19 | .21* | .15 | .35** | .53** | | |
| 7. Post-traumatic stress disorder (HTQ) | 1.72 | 0.49 | .20* | .13 | .41** | .26** | .55** | .52** | |
| 8. HRV (log RMSSD) | 3.52 | 0.56 | -.08 | -.21 | -.01 | -.14 | -.12 | -.10 | -.30* |

*Note.* The reported scales were derived from the following questionnaires: Harvard Trauma Questionnaire (HTQ); Perceived Ethnic Discrimination Questionnaire (PEDQ); Injustice Experience Questionnaire (IEQ); Beck Depression Inventory-II (BDI). The log-transformed (log) Root Mean Square of Successive Differences (RMSSD) was included as a measure of heart rate variability (HRV).
* p < .05; ** p < .01

### S2 Sample Distributions of Depression and PTSD Categories

| | Full Sample (*N* = 98) | | HRV Subsample (*n* = 64) | |
|---|---|---|---|---|
| Depression (BDI) | | + PTSD (HTQ ≥ 2.5) | | + PTSD (HTQ ≥ 2.5) |
| minimal (0-13) | 82 (83.67%) | 2 (2.04%) | 53 (82.81%) | 2 (3.13%) |
| mild (14-19) | 9 (9.18%) | 1 (1.02%) | 6 (9.38%) | 0 (0%) |
| moderate (20-28) | 6 (6.12%) | 3 (3.06%) | 4 (6.25%) | 2 (3.13%) |
| severe (29-63) | 1 (1.02%) | 1 (1.02%) | 1 (1.56%) | 1 (1.56%) |

*Note.* HRV = heart rate variability. Clinical categories of depression were assigned according to the Beck Depression Inventory-II (BDI). Post-Traumatic Stress Disorder (PTSD) was assessed with the Harvard Trauma Questionnaire (HTQ), which has a clinical cut-off value of ≥ 2.5.

### S3 Covariate Analysis for Age and Sex

| | *b* | *SE* | *z* | *p* | β |
|---|---|---|---|---|---|
| Model 1 | | | | | |
| PTSD ($R^2$ = .332) | | | | | |
| Trauma | 0.03 | 0.01 | 5.60 | < .001*** | 0.45 |
| Age | 0.01 | 0.01 | 1.64 | .102 | 0.14 |
| Sex (female) | 0.43 | 0.11 | 3.83 | < .001*** | 0.35 |
| Depression ($R^2$ = .243) | | | | | |
| Discrimination | 2.21 | 0.76 | 2.92 | .003** | 0.33 |
| Age | 0.43 | 0.13 | 3.31 | .001** | 0.30 |
| Sex (female) | 4.56 | 1.69 | 2.70 | .007** | 0.27 |
| PTSD ~~ Depression | 1.04 | 0.33 | 3.15 | .002** | 0.42 |
| Trauma ~~ Discrimination | 2.89 | 1.04 | 2.78 | .005** | 0.33 |
| Model 2a | | | | | |
| PTSD ($R^2$ = .428) | | | | | |
| Trauma | 0.02 | 0.01 | 3.89 | < .001*** | 0.36 |
| Perceived injustice | 0.02 | 0.01 | 3.97 | < .001*** | 0.38 |
| Age | 0.01 | 0.01 | 1.09 | .276 | 0.10 |
| Sex (female) | 0.28 | 0.11 | 2.49 | .013* | 0.23 |
| Depression ($R^2$ = .390) | | | | | |
| Discrimination | 1.49 | 0.69 | 2.16 | .030* | 0.22 |
| Perceived injustice | 0.30 | 0.09 | 3.41 | .001** | 0.39 |
| Age | 0.34 | 0.12 | 2.85 | .004** | 0.23 |
| Sex (female) | 2.62 | 1.50 | 1.75 | .080 | 0.15 |
| Perceived injustice ($R^2$ = .279) | | | | | |
| Discrimination | 3.76 | 0.87 | 4.32 | < .001*** | 0.42 |
| Age | 0.35 | 0.17 | 2.03 | .042* | 0.18 |
| Sex (female) | 6.82 | 2.11 | 3.23 | .001** | 0.30 |
| PTSD ~~ Depression | 0.66 | 0.26 | 2.54 | .011* | 0.32 |
| Trauma ~~ Discrimination | 2.89 | 1.00 | 2.80 | .004** | 0.33 |
| Indirect Effects on | | | | | |
| PTSD | 0.08 | 0.03 | 2.94 | .003** | 0.16 |
| Depression | 1.12 | 0.47 | 2.41 | .016* | 0.17 |
| Model 2b | | | | | |
| PTSD ($R^2$ = .390) | | | | | |
| Trauma | 0.03 | 0.01 | 4.41 | < .001*** | 0.45 |
| HRV | -0.23 | 0.08 | -2.95 | .003** | -0.25 |
| Age | 0.02 | 0.01 | 1.51 | .131 | 0.15 |
| Sex (female) | 0.39 | 0.14 | 2.90 | .004** | 0.28 |
| Depression ($R^2$ = .325) | | | | | |
| Discrimination | 3.15 | 1.05 | 3.02 | .003** | 0.46 |
| Age | 0.43 | 0.16 | 2.70 | .007** | 0.28 |
| Sex (female) | 3.90 | 2.34 | 1.67 | .100 | 0.19 |
| PTSD ~~ Depression | 0.87 | 0.41 | 2.12 | .034* | 0.36 |
| Trauma ~~ Discrimination | 2.82 | 1.46 | 1.93 | .054 | 0.31 |

*Note.* PTSD refers to Post-Traumatic Stress Disorder. HRV refers to heart rate variability and was measured as the log-transformed Root Mean Square of Successive Differences (RMSSD). ~~ refers to covariances. All parameters were obtained using bootstrapping with 1,000 iterations. Fit for all three models was insufficient: $\chi^2(6) = 15.50$, $p = .017$, CFI = 0.900, TLI = 0.766, RMSEA = 0.127 (90% CI [0.05, 0.207]) for Model 1; $\chi^2(7) = 18.63$, $p = .009$, CFI = 0.921, TLI = 0.774, RMSEA = 0.130 (90% CI [0.060, 0.204]) for Model 2a; $\chi^2(9) = 20.21$, $p = .017$, CFI = 0.846, TLI = 0.692, RMSEA = 0.139 (90% CI [0.056, 0.222]) for Model 2b.
* $p < .05$; ** $p < .01$; *** $p < .001$

**S4 Covariate Analysis for Migrants and Refugees**

| | *b* | *SE* | *z* | *p* | β |
|---|---|---|---|---|---|
| Model 1 | | | | | |
| PTSD ($R^2$ = .148) | | | | | |
| Trauma | 0.02 | 0.01 | 3.24 | .001** | 0.34 |
| Role (refugee) | 0.17 | 0.11 | 1.60 | .109 | 0.18 |
| Depression ($R^2$ = .097) | | | | | |
| Discrimination | 1.74 | 0.75 | 2.31 | .021* | 0.27 |
| Role (refugee) | 2.23 | 1.25 | 1.78 | .076 | 0.16 |
| PTSD ~~ Depression | 1.38 | 0.37 | 3.77 | < .001*** | 0.49 |
| Trauma ~~ Discrimination | 2.89 | 1.00 | 2.90 | .004** | 0.33 |
| Model 2a | | | | | |
| PTSD ($R^2$ = .343) | | | | | |
| Trauma | 0.01 | 0.01 | 2.39 | .014* | 0.26 |
| Perceived injustice | 0.02 | 0.01 | 5.09 | .024* | 0.46 |
| Role (refugee) | 0.11 | 0.10 | 1.13 | .109 | 0.12 |
| Depression ($R^2$ = .305) | | | | | |
| Discrimination | 1.10 | 0.70 | 1.57 | .118 | 0.17 |
| Perceived injustice | 0.35 | 0.09 | 3.81 | < .001*** | 0.46 |
| Role (refugee) | 0.88 | 1.21 | 0.73 | .464 | 0.06 |
| Perceived injustice ($R^2$ = .173) | | | | | |
| Discrimination | 3.22 | 0.94 | 3.42 | .001** | 0.37 |
| Role (refugee) | 3.57 | 1.73 | 2.07 | .039* | 0.19 |
| PTSD ~~ Depression | 0.77 | 0.25 | 3.02 | .003** | 0.35 |
| Trauma ~~ Discrimination | 2.89 | 1.00 | 2.90 | .004** | 0.33 |
| Indirect Effects on | | | | | |
| PTSD | 0.08 | 0.03 | 2.88 | .004** | 0.17 |
| Depression | 1.12 | 0.46 | 2.46 | .014* | 0.17 |
| Model 2b | | | | | |
| PTSD ($R^2$ = .250) | | | | | |
| Trauma | 0.02 | 0.01 | 2.85 | .004** | 0.39 |
| HRV | -0.24 | 0.08 | -2.97 | .003** | -0.27 |
| Role (refugee) | 0.11 | 0.13 | 0.85 | .395 | 0.11 |
| Depression ($R^2$ = .195) | | | | | |
| Discrimination | 2.72 | 1.07 | 2.55 | .011* | 0.41 |
| Role (refugee) | 2.49 | 1.53 | 1.63 | .103 | 0.17 |
| PTSD ~~ Depression | 1.20 | 0.47 | 2.59 | .010* | 0.43 |
| Trauma ~~ Discrimination | 2.82 | 1.46 | 1.93 | .054 | 0.31 |

*Note.* PTSD refers to Post-Traumatic Stress Disorder. Role refers to whether the participant belonged to the migrant or the refugee group. HRV refers to heart rate variability and was measured as the log-transformed Root Mean Square of Successive Differences (RMSSD). ~~ refers to covariances. All parameters were obtained using bootstrapping with 1,000 iterations. Fit for all three models was insufficient: $\chi^2(4) = 32.42$, $p < .001$, CFI = 0.703, TLI = 0.256, RMSEA = 0.269 (90% CI [0.188, 0.359]) for Model 1; $\chi^2(5) = 31.38$, $p < .001$, CFI = 0.817, TLI = 0.451, RMSEA = 0.232 (90% CI [0.158, 0.313]) for Model 2a; $\chi^2(7) = 29.88$, $p < .001$, CFI = 0.693, TLI = 0.387, RMSEA = 0.226 (90% CI [0.146, 0.312]) for Model 2b.

* $p < .05$; ** $p < .01$; *** $p < .001$
